# Experiences of staff providing specialist palliative care during COVID-19 *A multiple qualitative case study*

**DOI:** 10.1101/2021.11.17.21266437

**Authors:** Andy Bradshaw, Lesley Dunleavy, Ian Garner, Nancy Preston, Sabrina Bajwah, Rachel Cripps, Lorna K Fraser, Matthew Maddocks, Mevhibe Hocaoglu, Fliss EM Murtagh, Adejoke O Oluyase, Katherine E Sleeman, Irene J Higginson, Catherine Walshe, on behalf of the CovPall study team

## Abstract

**Objectives:** To explore the experiences of, and impact on, staff working in palliative care during the COVID-19 pandemic.

**Design:** Qualitative multiple case study using semi-structured interviews between November 2020 and April 2021 as part of the CovPall study. Data were analysed using thematic framework analysis.

**Setting:** Organisations providing specialist palliative services in any setting.

**Participants:** Staff working in specialist palliative care, purposefully sampled by the criteria of role, care setting and COVID-19 experience.

**Main outcome measures:** Experiences of working in palliative care during the COVID-19 pandemic.

**Results:** Five cases and 24 participants were recruited (n=12 nurses, 4 clinical managers, 4 doctors, 2 senior managers, 1 healthcare assistant, 1 allied healthcare professional). Central themes demonstrate how infection control constraints prohibited and diluted participants’ ability to provide care that reflected their core values, resulting in experiences of moral distress. Despite organisational, team, and individual support strategies, continually managing these constraints led to a ‘crescendo effect’ in which the impacts of moral distress accumulated over time, sometimes leading to burnout. Solidarity with colleagues and making a valued contribution provided ‘moral comfort’ for some.

**Conclusions:** This study provides a unique insight into why and how healthcare staff have experienced moral distress during the pandemic, and how organisations have responded. Despite their experience of dealing with death and dying, the mental health and well-being of palliative care staff was affected by the pandemic. Organisational, structural, and policy changes are urgently required to mitigate and manage these impacts.

## Background

COVID-19 has additionally stressed already stretched healthcare systems,. ^1-3^, influencing how organisations, and professionals that work within them, are able to respond to patient and carer needs. A combination of dealing with death and dying, risks of infection, personal loss/grief, and operating in insufficiently resourced services has resulted in many experiencing anxiety, depression, insomnia, burnout, and post-traumatic stress disorder. ^4-8^

Palliative care is a unique speciality in that staff are used to dealing with dying and may have been less affected by this aspect of the COVID-19 pandemic. Nevertheless, in responding to COVID-19, palliative care professionals have been confronted with constraints and obstacles (e.g., making complex and difficult decisions, infection control, dealing with uncertainty, and recognising deep inequities ^6, 9, 10^) that have severely challenged their ability to provide care in accordance with their professional values. These values include alleviating suffering and enhancing the quality of life of dying patients and their families through the adoption of a holistic, compassionate, person-centred, dignified, safe, and multidisciplinary approach. ^11^

Consequently, understanding how palliative care professionals, who choose to work with those who are dying, responded to the pandemic is key. It is important to understand how individual, organisational, and policy-based changes can be made to alleviate and manage the impact of the pandemic on staff. ^12^ The aim of this study, therefore, was to explore the experiences of, and impact on, palliative care staff working during the COVID-19 pandemic to illuminate both their experiences and how this may help an understanding of supporting healthcare staff and organisations more generally.

## Methods

A descriptive qualitative multiple case study, ^13, 14^ part of the ‘CovPall study’; a project aiming to understand the multinational response of specialist palliative and hospice care services to the COVID-19 pandemic. ^5, 6, 8, 15, 16^ It was guided by the following research questions:

- How has the COVID-19 pandemic impacted staff working in palliative care?
- How did organisations respond to the impact of the COVID-19 pandemic on staff well-being?

### Case definition, selection, and recruitment

Cases were defined as organisations providing specialist palliative care services across any setting. Potential sites that met the inclusion/exclusion criteria were identified from responses to an initial CovPall survey, ^5^ with cases sampled for maximum variability against key criteria until sufficient organisations were recruited (Table 1).

**Table 1:** Inclusion and exclusion criteria for the recruitment of case study sites and participants

| **Cases** |  |
| --- | --- |
| **Inclusion criteria** | Organisation providing specialist palliative care services.<br>Respondent to CovPall survey, with agreement for further contact.<br>Restricted to English Hospice organisation respondents due to constraints of research approvals. |
|  | Sampled against criteria to maximise variability:<br>Number of services and setting types that organisations provided<br>Whether adult and/or paediatric services were provided<br>Experience of providing care to those with COVID-19<br>Whether minority ethnic populations were served<br>Variability in their initial service changes to COVID-19 |
| **Participants within selected cases** |  |
| **Inclusion criteria** | Working or volunteering within the chosen case and able to provide rich data on the experience of care provision during COVID-19 (e.g. senior managers, clinical managers, direct healthcare staff).<br>Aged 18+ |
| **Exclusion criteria** | Patients known to the service, or their family carers. |

### Within case participant selection and recruitment

Key contacts within each case study site identified potential participants who met the inclusion criteria (Table 1), purposively sampled to reflect variations in professional role, work setting, and experience in responding to COVID-19. Key contacts distributed study information (participant information sheets and consent forms) to those who could provide rich insight into the aims of the study.

### Theoretical propositions

In line with case study research strategies, ^13^ we used the survey data to develop initial theoretical propositions to guide data collection and analysis:

1. The type of service provider organisation made a difference to the way that specialist palliative care responded to COVID-19.
2. The context within which the service provider organisation operated affected their response. This may include geography (e.g., when they first experienced COVID-19, local healthcare organisational factors) and factors known to affect service use (e.g., deprivation, ethnicity).
3. Exposure to COVID-19 patients (e.g., numbers of patients, and whether patient were dying with or from COVIID-19 or other diseases) made a difference to the service response to COVID-19.
4. Systems or processes that supported responsive decision-making affected response to COVID-19 which included aspects of integration with other services and organisational leadership.

### Data collection

Single online (via Microsoft teams) or telephone semi-structured interviews were conducted. The interview guide (eTable 1) was iteratively developed throughout the study. Participants were asked to reflect on how they had experienced working throughout the COVID-19 pandemic, how they felt their organisation had responded to challenges during this time, and ways in which we could learn from the pandemic to inform future practice. Interviews were conducted by AB (male, research fellow, PhD) and IG (male, research fellow, PhD), both of whom had previous interviewing experience. They were digitally recorded, anonymised, and transcribed verbatim. Field notes were made during and after each interview. On average, interviews lasted 39 minutes (range 22-80 minutes). Data were collected between November 2020 and April 2021. This coincided within (September 2020 - January 2021) and after the second wave of the COVID-19 pandemic in England.

### Data analysis

Thematic framework analysis was used to analyse data. ^17^ This approach allowed us to conduct within- and between-case pattern matching, thus enabling a process in which we could identify and explore where participant responses converged/diverged, and how this may have been affected by different contextual factors. ^17, 18^ This approach involved constructing themes through five interconnected stages: (i) familiarisation; (ii) coding transcripts to construct an initial analytic framework; (iii) indexing and further refinement of the analytic framework; (iv) charting; (v) mapping and interpreting the data theory/theoretical concepts to make sense of and explain our data. Data were initially analysed within cases and then between cases.

Moral distress was identified as a useful lens through which these data could be viewed. Generally, moral distress refers to ‘the experience of being seriously compromised as a moral agent in practicing in accordance with accepted professional values and standards.’ ^19^ It has historically focused on institutional/organisational obstacles that impact healthcare professionals’ ability to deliver care in accordance with their values. ^9, 10^ Recent literature, however, has recognised the importance of appreciating sources of moral distress that derive from ‘broad[er] challenges of the health services system’, ^12^ incorporating regional, national, and global issues. ^20^ We adopt the latter perspective when referring to moral distress throughout this paper.

The analysis process was primarily conducted by AB, LD, and IG. Throughout this process, co-authors CW and NP (and the wider CovPall team) acted as ‘critical friends’. ^21^ This was through cross-checking coding, and discussing, debating, and providing alternative interpretations of data until the research team were happy that interpretations of data accurately reflected participant accounts.

### Ethics committee and other approvals and registrations

Research ethics committee approval was obtained from King’s College London Research Ethics Committee (21/04/2020, Reference; LRS19/20-18541), with additional local approval from Lancaster University FHMREC 24.11.2020 Reference FHMREC20057). The study was registered on the ISRCTN registry (27/07/2020, ISRCTN16561225) and reported in line with the COREQ checklist. ^22^

### Findings

Five cases drawing from the experiences of 24 participants were included (Table 2). The findings are presented as a cross case analysis and are represented as four themes and two subthemes (see figure 1). Additional example quotes for each theme and sub-theme are in supplementary materials (eTable 2).

**Table 2:** Case and participant characteristics

|  | Case one | Case two | Case three | Case four | Case five |
| --- | --- | --- | --- | --- | --- |
| **Case characteristics** |  |  |  |  |  |
| Region | North West England | South East England | North West England | North East England | South East England |
| % NHS funding | Approx. 35 % | Approx. 25% | Approx. 30 % | Approx. 25 % | Approx. 40 % |
| Patients served | Adult | Adult/children | Adult/children | Adult | Adult |
| Services provided | Inpatient palliative care unit, home palliative care team, home nursing | Inpatient palliative care unit, home palliative care team, home nursing | Inpatient palliative care unit, hospital palliative care team, home palliative care team, home nursing | Inpatient palliative care unit, hospital palliative care team | Inpatient palliative care unit, hospital palliative care team, home palliative care team, home nursing |
| Population served* | Urban, suburban and rural<br>Primarily White | Urban, suburban and rural<br>Primarily White | Suburban and rural.<br>Primarily White. | Suburban and rural.<br>Primarily White | Urban and suburban.<br>Primarily White |
| **Study participant characteristics** |  |  |  |  |  |
| Data collected | 11/20-04/21 | 11/20-03/21 | 12/20-01/21 | 03/21 | 04/21 |
| COVID wave (UK)† | Wave 2/post wave 2 | Wave 2/post wave 2 | Wave 2 | Post wave 2 | Post wave 2 |
| Professional role | Nurse n= 4<br>Clinical manager n = 1<br>Doctor n= 1 | Nurse n= 3<br>Clinical manager n=2<br>Health care assistant n= 1 | Nurse n= 2<br>Senior manager n =1<br>Doctor n= 3 | Nurse n=1<br>Clinical Manager n= 1<br>Allied Health Care Professional n=1 | Nurse n=2<br>Senior manager n=1 |
| Ethnicity | White n= 6 | White n= 5<br>Asian/Asian British=1 | White n= 6 | White n= 3 | White n= 2<br>Missing =1 |
| Time worked in palliative care (months)<br>mean/range | 98 months (48-360) | 108 months (36-192)<br>Missing=1 | 109 months (18-180) | 196 months (60-360) | 192 months (360-24)<br>Missing=1 |
| Time worked in current position (months)<br>mean/range | 93 months (12-240) | 14.8 ? 15 months (3-36)<br>Missing=1 | 61.3 months ? 61(18-120) | 72 months (60-96) | 78 months (132-24)<br>Missing=1 |

**Figure 1:**
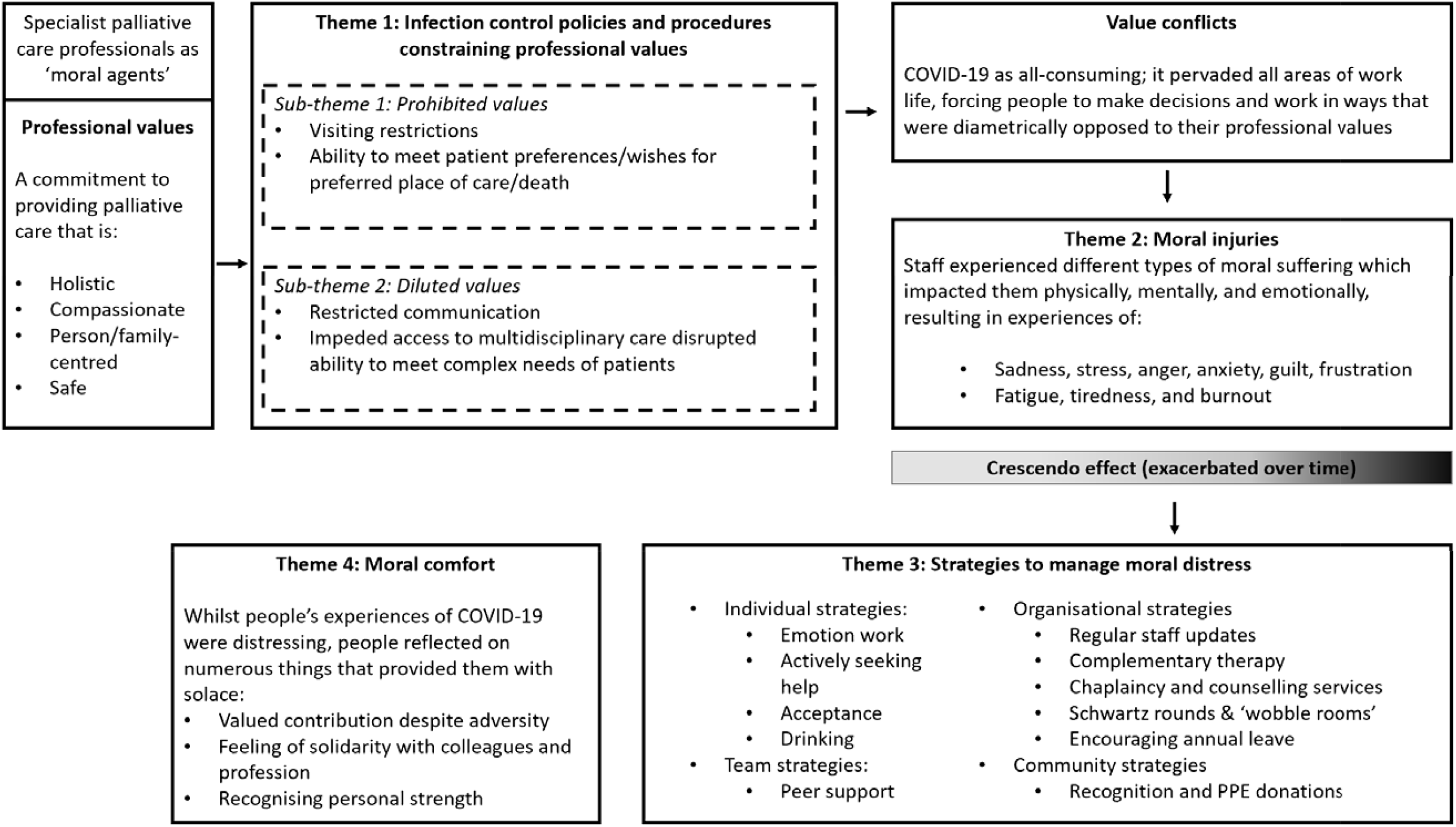
An overview of the themes and sub-themes that represent the processes through which participants in this case study experienced moral distress

### Theme 1: Infection control constraining professional values

The most common constraints to practicing in line with professional values were directly or indirectly related to infection control policies/procedures. These constraints triggered moral distress by either prohibiting or diluting the abilities of individuals and organisations to uphold and practice in accordance with their professional values. A unifying pattern across the cases was that the root cause of moral distress was not primarily the result of looking after patients who were dying, but because of care constraints impacting on *how* they were able to care for dying patients.

#### Sub-theme 1: Prohibited values

In some instances, the impacts of infection control procedures prohibited staff’s ability to provide care in accordance with their professional values. In particular, restricted visiting policies forced participants to make decisions and operate in ways that were opposed to the holistic and person/family-centred values of palliative care. In the hospital setting, staff had to inform families that no visiting was allowed (even at the end of life) whereas in the hospice settings only a limited number of visitors were generally permitted. Witnessing patients die without loved one’s present, alongside having to deal with the conflicts that visiting restrictions caused was particularly distressing:

> ***‘****Throughout this whole Covid experience, what stays with me the most are those conversations with loved ones and family members to say: ‘I am really sorry, we can’t enable a visit’, or if you do it is a one-off kind of hour visit… they have been some of the hardest conversations that I have had in my whole nursing career … you can’t help but feel that you have not done enough, even though I know that we have … it just goes against the grain of everything we do’ participant 5, case 1, nurse*

Visiting restrictions also impacted staff’s ability to visit patients’ homes. As referrals increased in the community, staff were required to triage who did and did not require an in-person visit to reduce the risk of infection. Consequently, some participants felt that the care they were providing was different/inadequate and compromised compared to before COVID-19. Feeling care was compromised, as well as managing disagreements with family carers over whether an in-person visit was necessary, was a source of moral distress for some:

> *‘we cut down the visits we were doing, so in the home care team the visits would be done if they really needed to… But, anybody where we could do it over the phone, because you were just minimising contact and obviously reducing the risk of spreading the virus. But, I think some family members did see that as ‘but you are not really here, you are not coming out and doing visits, you are just over the phone’ … it is trying to find a … tactical way of saying that there is no need to increase that risk for something that can be done over the phone. participant 3, case two, nurse*

Infection control issues also prohibited staff’s capacity to provide care that was aligned with patient preferences. Not being able to admit patients requiring aerosol generating procedures into hospice inpatient units, or an inability to discharge patients out of hospital or hospice, placed staff in situations where they were sometimes unable to honour peoples wishes regarding preferred place of care/death:

> *‘not being able to get the patients out of hospital because care homes won’t accept COVID-positive patients. …. people who don’t have long left to live and don’t want to die in hospital, you know, delaying that, there’s more chance that they are going to die in hospital if we can’t get them out. It’s been one of the biggest challenges, discharge, it’s so difficult to juggle on a daily basis. The number of beds, patients coming in, trying to get patients out, it’s horrendous*.*’ Participant 3, case three, doctor*

#### Sub-theme 2: Diluted values

In some situations, whilst staff were able to carry on providing palliative care within infection control constraints, they recognised it diluted their ability to provide care in line with their professional values. Many raised concerns about how their ability to care for patients and families with the same level of compassion and empathy as prior to the pandemic was constrained by visiting restrictions, social distancing, and unprecedented staff shortages. Sensitive conversations, such as breaking bad news or GP verification of death were carried out remotely, whilst in-person communication was impeded by Personal Protective Equipment. Being unable to draw on non-verbal communication skills and visual cues made care feel physically and emotionally detached, undermining practitioners’ capacity to develop relationships, fully support, and comfort patients and carers at profoundly important moments. This posed a moral dilemma for staff; whilst many participants recognised the necessity of these safety measures, witnessing and managing the suffering and pain that they caused families and patients was deeply distressing:

> *‘PPE is just such a barrier between us and the patients…. it’s a bit more impersonal. Obviously, we deal with patients and their families that are dying, and often patients and family, they’re quite emotional, and we can sort of maybe just sort of put our arm round them or embrace them in some way, which is something we can’t do at the moment … And it is harder for us, because obviously we do this job because it’s a very rewarding job to do, and so I think it is different for us, not being able to comfort somebody*.*’ participant 4, case two, health care assistant*

Infection control policies also impeded access to the wider multi-disciplinary team and diluted the level of support they were able to provide. In some cases, this was due to services being suspended, adapted, or provided remotely, or staff and volunteers having to self-isolate or shield. This led to moral distress as staff were concerned that patients with complex needs were not receiving the level of support they required:

> *‘the other big thing that the staff have been seriously challenged with is their professional values of very comprehensive holistic patient-centred care that is the hallmark of good palliative care and* … *so many restrictions have had to be put in place and the services that we’ve had to suspend really and perhaps day surgery [therapy] or complementary services, things have had to go to remote conversations and consultations. They’ve found it very, very difficult to accept that change in standards or those constraints to being able to get that high standard of personal care*.*’ participant 1, case three, doctor*

### Theme 2: Moral injuries

At the beginning of the pandemic, clinicians reported feelings of anxiety/fear due to dealing with an unknown disease and new infection control procedures. As more was known about COVID-19 and access to PPE improved, participants reported that they generally became less fearful and worried. Instead, these feelings were replaced by those of sadness, stress, anger, guilt, frustration, and fatigue as a result of repeatedly experiencing scenarios in which their professional values were challenged. These responses represented ‘moral injuries’ and exemplified how moral distress had profound impacts on the physical, mental, and emotional health of staff across cases, settings, and roles:

> *‘some days I have really struggled – I am not going to lie. I have absolutely sobbed my heart out, thinking about stuff that I have gone through and seen and conversations that I have had to have with family members. But, ultimately you go back into work the next day and you carry on, because you know that you have to because you have got a job to do, and there are patients and people there that are relying on you to do that, do you know what I mean? So, yes it has been … it has been challenging, mentally and physically*.*’ participant 5, case one, nurse*

Whilst experiences of moral injuries were similar across cases, the source of moral distress was sometimes role dependent. Whilst policies around infection control were often the source of moral distress for clinicians providing direct patient care, those in managerial positions had to make difficult decisions on suspending/reducing services, furloughing staff, and/or making redundancies (case one and three) because of reduced income. They also worried about and felt responsible for their staff’s wellbeing and safety:

> *‘when I look back on it now, really quite - difficult’s the wrong word - but conversations with colleagues where we were basically discussing the ethics of putting our staff in front of patients with COVID knowing that they might catch it and they might die from it and that was really hard. We were asking them to do superhuman things*.*’* participant 1, case 5, senior manager

Across cases, a ‘crescendo effect’ occurred in which the effects of moral distress accumulated and escalated progressively over time. This was likened to ‘*a drip, drip effect’* [*participant 5, case 1, nurse]* and explained how tiredness, fatigue, and frustration affected team dynamics and, in some cases, led to or exacerbated staff conflicts. Moreover, it also exemplifies the process through which some staff became burnt out which, in worst case scenarios, led to staff leaving their roles:

> *‘when wave two hit, there was a real oh my God can we do this again? I think it is that whole thing – you didn’t have any of the fight that you had the first time – it was a case of right come on, we have got to do it, but it has definitely been done very well, but it is hard. It is more of a slog this time than it was the first time… I think the actual day-to-day care wasn’t more difficult, I think people were more tired. And, I think the fact that the impact it has had on people, on staff, externally so your whole lifestyle – people haven’t got that … same resilience I don’t think, from the first wave*.*’ Participant 2, case one, clinical manager*

Laced throughout some participant accounts was a sense that they perceived themselves to be relatively powerless in addressing the fundamental causes of moral distress:

> *‘ultimately the saddest thing about it all is that really there isn’t anything that we can do to take that away – this is the situation that we are in and it is awful and it is horrible, and people are struggling with it up and down the country, and all you can do at times is just let somebody talk or just let somebody get upset or get angry. ‘ participant 5, case one, nurse*

### Theme 3: Strategies to manage moral distress

The detrimental impacts of moral distress were recognised early, and a variety of individual, team, and organisational strategies were used to help manage its effects. At an individual level, participants undertook emotion work and adopted their own strategies to manage their moral distress. This could include less healthy strategies (such as drinking alcohol more heavily), but also strategies such as accepting their situation, embracing the normality of work, actively seeking help, and empathising with patients and families:

> *‘I think my mental health has deteriorated but I think everyone’s has so I think that’s fine. I definitely reached a point where I thought, “I’m drinking too much” because it became*… *When you’re at home and you’re stressed you’re like, “What can I do? I can’t go to the gym, I could go out for a run but it’s dark and I don’t want to be murdered so I’m going to have a glass of wine”. And then you have one glass of wine and you’re like, “Oh that does feel better. If I have another one that’ll make me feel even better … And then the next day I’m like, “I’m not going to drink today” then I have a really stressful meeting and I’m like, “No, I am, I’m going to have a drink tonight”… But yeah, so mental health, definitely, weight, alcohol dependency’ participant 5, case two, nurse*
>
> *‘And just tend to sit and cry with relatives… on the one hand it’s not really the done thing, but on the other hand I guess it shows that you’re human and it shows that you are absorbing some of the impact of that emotional situation. And it’s showing that you kind of respect that it is so sad*.*’ participant 2, case four, nurse*

At a team level, participants noted the value of peer support in helping them to manage moral distress. Moreover, across cases, participants felt organisations did the best they could to support staff in very difficult circumstances through providing regular staff updates, ‘wobble rooms’, access to patient therapy/support services, Schwartz rounds, and encouraging leave. There were some concerns that staff did not always have the time to access support and strategies that required staff to be on site were not accessible to all:

> *‘They created a wobble room for people to go and wobble in, it’s difficult again though with everybody off site now and working from home I think for me anyway personally the main impact of that wobble room is just knowing that they’ve thought about it that’s reassuring that they’re mindful of our mental health and our emotional needs but it’s not actually in practice that useful because nobody… especially for the community staff, they don’t get that*.*’ participant 3, case one, nurse*

On a practical level, ensuring the hospice had adequate supplies of PPE was important to reassure staff. The wider community donated gifts and supplies of PPE and food so staff *‘knew that people out there were still thinking about us*.*’* [Participant 1, case four, clinical manager].

### Theme 4: Moral comfort

Despite the impacts of moral distress, some participants spoke about how they experienced comfort and solace in their situation as they felt they were making a valued contribution to the pandemic response. Staff also recognised their own personal strength and how solidarity with colleagues was developed or strengthened in responding to the pandemic:

> *What we learnt as a service was learnt that we are a good team, that we can respond, that we’re respected and valuable members of our local health and social care system and that we can add real value to that, that certainly as a management team we’ve been able to be very flexible and adapt very quickly and move people around the service and that we’ve been able to reach more people and keep our education going virtually, that we’ve been able to still have a big impact and, you know, without undermining the quality of the care that we give too much…. when we look back on this what will we be proud of in terms of what was our contribution*.*’ participant 1, case three, doctor*

## Discussion

By using palliative care as a clinical exemplar, this study highlights how staff working across healthcare settings are likely to have been affected by the pandemic, alongside lessons that can be learnt about how moral distress can be prevented, alleviated, or mitigated. Constraints related to COVID-19 infection control policies and practices were central to experiences of moral distress by prohibiting and/or diluting staff’s capacity to provide care that was aligned to their professional caring values. Experiences of moral distress had a detrimental impact on the well-being of staff by causing ‘moral injuries’ in which participants experienced feelings of sadness, stress, anger, guilt, frustration, and fatigue. These feelings crescendoed over time whereby the impacts of moral distress had a cumulative effect that worsened as the pandemic progressed. Various individual, team, organisational, and community strategies were drawn on to address the impacts of moral distress (see Figure 1), and despite working through adversity, some participants reported feelings of ‘moral comfort’ by making valued contributions in response to the pandemic. The final theoretical propositions were elaborated as:

1. All organisations recognised the risks of moral distress and responded in similar ways.
2. Whilst experiences and signs of moral distress were similar across cases, settings and participants, the sources of moral distress were setting and role dependent.
3. As the length of the pandemic continued, the impacts of moral distress progressively accumulated and worsened for some.
4. Despite the accumulation of moral distress, some staff experienced a sense of comfort and solace because they felt they were making a valued contribution to the pandemic response.

Fundamental to staff’s experiences of moral distress was a sense of discordance between wanting to deliver care in specific ways, but not being able to. Whilst some constraints that contributed to moral distress were COVID-specific (i.e., infection control policies), many (such as decision-making conflicts, insufficient resources, staff shortages, funding issues, and patient complexity) already existed prior to the pandemic. ^23-27^ This aligns with evidence demonstrating how many healthcare staff already experienced moral distress/injuries prior to COVID-19, but how the pandemic has brought these phenomenon into sharper focus. ^27^ The increased risk of moral distress for health care staff during the pandemic has been acknowledged by regulatory bodies and governments internationally, ^27-29^ and this concern is supported by emerging evidence in the fields of acute care, ^30^ community care, ^31^ intensive care, ^32^ medical family therapists, ^33^ mental health, ^34^ and medicine more generally. ^10^ Compared to many of these specialities, due to their specialist training and knowledge, palliative care staff may have been expected to be better prepared to manage experiences of death and dying on the scale seen during the COVID-19 pandemic. That many staff within palliative care experienced moral distress in witnessing *how* people died, there is a likelihood of even more profound distress, stress, and burnout in generalist staff who – alongside dealing with structural and policy constraints of COVID-19 – were exposed to death and dying on a scale unimaginable to most healthcare professionals outside of the pandemic.

The detrimental impact of moral distress on staff well-being aligns with literature demonstrating how repeatedly occupying spaces of moral distress can negatively affect the physical, mental, and emotional health of healthcare workers. ^35-41^ These findings support and build on emerging evidence of the impact of COVID-19 on staff within ^42-44^ and outside ^45-49^ of palliative care by providing detailed insights into how and why staff experienced moral distress throughout the pandemic. If the impacts of moral distress are sustained without being recognised, prevented, or dealt with appropriately, it can decrease the capacity of health professionals to deliver high quality care, lead to burnout, and increase the likelihood of staff making errors and leaving roles. ^10, 25, 50, 51^ Considering there are already high levels of burnout and staff shortages in many healthcare settings, with shortages projected to worsen by 2030, ^29, 52, 53^ retention of skilled personnel is crucial. This is so that healthcare systems retain the capacity to meet projected increases in global demand/need for palliative care ^54, 55^ and across all healthcare sectors more generally. ^29, 56, 57^ Therefore, understanding what changes can be made to prevent, alleviate and manage the short and long-term impacts of moral distress on all healthcare staff - both throughout and after the pandemic - is crucial to the future provision of healthcare. ^12^

In effectively preventing and mitigating moral distress across healthcare settings, interventions need to be targeted at multiple levels of practice (individual, interpersonal, organizational, and policy-levels). ^20^ Some evidence suggests that proactive individual and interpersonal strategies to manage moral distress may be learned through experience, ^58, 59^ and examples of these during COVID-19 are demonstrated within this and other studies. ^45, 60^ However, strategies to manage moral distress should not solely be placed on individuals; governments and organisations have a duty of care to healthcare staff, and it is important that they bear responsibility in developing structures and processes of care that address the causes of moral distress in order to facilitate staff well-being and prevent and/or mitigate workforce shortages. ^51^ Accordingly, Rodney ^20^ proposes the adoption of a relational ethical lens in managing moral distress whereby underpinning any intervention is an appreciation of the interconnectedness of people and structures. Supporting any individual or team level strategies to mitigate moral distress, therefore, should be national policy and organisational level solutions that create environments where staff feel supported and capable in delivering care. The British Medical Association propose numerous structural solutions that government and institutions may consider in achieving this. These include ensuring adequate funding and resourcing, increasing staffing, empowering doctors, developing an open and sharing workplace culture, providing organisational support to staff, and streamlining bureaucracy. ^27^ Potentially useful interventions may include Schwartz rounds, attention to staffing levels, and flexible working policies. ^61^ Future research on how to best achieve these solutions, alongside how organisations can ensure that they are accessible to staff across all roles and settings of care (including remotely), is needed.

A strength of this study lies in the adoption of a case study research design. This assisted us in providing rich and detailed insights into the processes through which responding to the COVID-19 pandemic impacted staff working in real-life clinical settings. ^62^ Through purposefully sampling cases and participants, using theoretical propositions, and constructing thick descriptions of findings and methods, we propose that ‘naturalistic generalisations’ may be made through findings resonating with healthcare staff within and outside of palliative care. ^63^ ‘Analytic generalisations’ may also be made through demonstrating the applicability and value of moral distress as a concept to understand healthcare staff’s experiences of responding to the pandemic. ^13, 63^ A limitation of this study, however, is that it relied on single individual interviews collected at only one timepoint. Whilst these provide a snapshot in which participants could retrospectively reflect on the impact of COVID-19, the long-term impact of COVID-19 on staff, alongside the sustainability/effectiveness of organisational responses, is not clear. Further longitudinal work that addresses these gaps will be a useful addition to the literature. Moreover, these data represent staff experiences of responding to COVID-19 from within a particular sector, and whilst there is likely to be overlap in experiences between healthcare settings, the nuances in experiences across other healthcare contexts (e.g., the public and private sectors) is not captured.

## Paragraph 6: Conclusion

Despite their experience of dealing with death and dying, the mental health and well-being of palliative care staff was affected by the pandemic. Key findings demonstrated how infection control constraints prohibited and diluted participants’ ability to provide care that reflected their core values, causing moral distress. Despite feeling some sense of comfort through contributing to the pandemic response, and although different strategies were used to manage moral distress, the impacts of the COVID-19 pandemic on staff well-being progressively worsened over time. Organisational, structural, and policy changes are urgently required to mitigate and manage these impacts to ensure quality of care and retention of staff.

## Data Availability

Applications for use of the survey data can be made for up to 10 years, and will be considered on a case by case basis on receipt of a methodological sound proposal to achieve aims in line with the original protocol. The study protocol is available on request. All requests for data access should be addressed to the Chief Investigator via the details on the CovPall website (https://www.kcl.ac.uk/cicelysaunders/research/evaluating/covpallstudy, and) and will be reviewed by the Study Steering Group.

## Acknowledgements

This study was part of CovPall, a multi-national study, supported by the Medical Research Council, National Institute for Health Research Applied Research Collaboration South London and Cicely Saunders International. We thank all collaborators and advisors. We thank all participants, partners, PPI members and our Study Steering Group. We gratefully acknowledge technical assistance from the Precision Health Informatics Data Lab group (https://phidatalab.org) at National Institute for Health Research (NIHR) Biomedical Research Centre at South London and Maudsley NHS Foundation Trust and King’s College London for the use of REDCap for data capture.

