## Supplementary File 1 for "Experiences of staff providing specialist palliative care during COVID-19 *A multiple qualitative case study*"

**eTable 1: Interview guide**

| **Question 1** | What is your role/duties within your specialist palliative care service? |
| --- | --- |
| **Question 2** | How do you feel your organisation has responded to the COVID 19 pandemic? |
| **Question 3** | What opportunities, if any, has the pandemic brought for your service?   I. Impact of opportunities on patients and family carers II. Impact of opportunities on clinical/non-clinical staff III. Facilitators and barriers to identifying/responding to opportunities. |
| **Question 4** | What do you think have been some of the main challenges of running your service during the COVID-19 pandemic?   I. What was it about these issues that made adjusting to them so challenging? II. How has this impacted on patients and family carers? III. Impact of challenges on clinical/non-clinical staff? |
| **Question 5** | Can you tell me a bit more about some of the innovations and changes in practice made in response to some of these issues? For example:   I. What innovations and changes do you think were particularly effective in achieving what you wanted it to? What do you think helped in implementing these changes? II. Alternately, what changes were made that are no longer practiced? And why do you think these changes were not effective, especially when compared to changes that were highly effective? III. If there was something you could alter in relation to the changes in practice to make it more effective, what would it be? |
| **Question 6** | What changes that have been made in your setting do you think would be effective in other palliative care settings. Why? |

**eTable 2: Additional quotes for each theme and sub-theme**

| **Theme** | *Sub-theme* | **Example quotes from participants** |
| --- | --- | --- |
| **Infection control constraining professional values** | *Prohibited values* | *‘I think we tried to facilitate as much as we could kind of remote contact so through FaceTime, that kind of thing, phone calls. And, again, that depends on that patient’s ability to be able to engage in that. But just holding a phone up to somebody’s ear while somebody said goodbye to them down the phone and things like that. Monumentally distressing … you nurse in a hospice because you want to give that kind of gold standard of care and it’s felt at time difficult to do that because we’ve not been as holistic as we ordinarily would be because of the kind of constraints around visiting and things like that.’ participant 2, case four, nurse* |
|  | *Diluted values* | **‘***normally I would be getting patients out of the rooms and into corridors, walking more and I have a physiotherapy room which I haven't been able to bring patients down to so they have been confined to the rooms so they've been more deconditioned than normal, I've not been able to do the same amount of rehab. I have a set of steps in the therapy room so I can't bring patients in here because I can't move them around, can't move patients around the ward and also the extra cleaning and like many therapy rooms across the country, have turned into PPE rooms because I'm sat in here now and there must be about 20 boxes of PPE.’ participant 3 case four, Allied health care professional* |
| **Moral injuries** |  | *‘one of other big challenges is we lost pretty much all our volunteers that were supporting all of this because many of them were in their 60s and 70s and when you take Covid age into account with multiple comorbidities quite a lot their Covid ages went above 70, and at that point people still didn’t know how dangerous this was so it was like, “Sorry, guys, you’ll have to go home”, and then of course that then has impacts on their emotional health so we then created virtual networks and friends groups and things like that but it’s not the same as their feelings of self-worth when wherever they volunteered they were feeling they were making a difference.’* participant 2, case three, CEO  *‘You have to think about properly what you're doing and you're exhausted. It’s hard. … you do, you feel kind of guilty in a way or you feel pressured, you think, “God, you know, if it [COVID-19] spreads then it’s going to be my fault”… I was definitely worried about it… One of the male bay did end up, all of them in the bay ended up testing positive and obviously we don’t know how that happened … I know a lot of people on the team were really, really sort of upset about it and anxious and things being, you know, was it me that give it them?’* participant 6, case one, nurse  *‘we’ve had to have redundancies … even though it’s a very well-resourced hospice and very well managed in terms of reserves and things… so that’s not a positive obviously … there’s no way of managing it well, is there, really, it’s not great… the biggest stress for people is not having a job and not being able to maintain their family and themselves and stuff, so it’s not easy at all … you’re having to make people redundant, it’s awful.’ participant 20, case 3, doctor*  *‘I think everybody’s quite at this point now, I mean god we’re nearly a year into it, everybody’s a bit fractious and a bit sensitive and a bit tired and do you know what I mean, picking up on everything that other people say and that’s quite difficult… everybody’s just tired, anxious, Covid-ed out with it all …it’s been a really long anxious time and I never would have thought it was going to go on this long … people are a little bit twitchy and a little bit short with each other and it’s about just mustering up that last bit of patience and trying to keep the team going’ participant 6, case three, nurse*  *‘I think I felt more the second wave than the first wave. And when I had – I kind of resignation within myself and I was saying every time this needs to be done, it won’t be forever. On the second one I was like, “Oh God, here we go again’ participant 3, case five, nurse* |
| **Strategies to manage moral distress** |  | **Individual strategies**  *‘I was just saying to kind of anyone them like mental health advice lines and your colleagues and everyone are there for you so just take advantage of them because there’s no – I mean like I say the job is hard enough but especially when you're in lockdown and you can’t even go to the gym, you can’t see your friends, you can’t, you know, do things that would normally feel benefit… I would definitely advise everyone to kind of seek, you know, some kind of advice or help or just speak to people that you can speak to and trust and feel comfortable with because that’s what got me through the last 12 months.’* participant 6, case one, nurse  *‘I think second lockdown we were more used to it. You know, it was sort of like a, yeah, well we’ll just have to go through it again. We just accepted it.’* participant 1, case four, clinical manager  **Team strategies**  *‘I also think we have got a fantastic team, even prior to Covid… that support from your colleagues is vital really, because ultimately you can come home and you can … you can say to your loved ones you have had a bad day, but nobody really kind of understands what you have gone through that day, other than the people that are dealing with it as well, so I think teamwork has been a massive, massive factor for us, in just kind of supporting each other.’* participant 5, case 1 , nurse  *‘we seem to be a very close-knit team now which is really nice … You just try to listen to each other and we could tell when someone was down, so we were there for each other. And I would say that, yeah, probably nine out of ten times we would know what the other one is going through or feeling. It’s not always the same. I understand that. But most of the time we were trying to predict how someone else is feeling or what their day is going to be like. So we just be there, extra motivating, encouraging and just a little bit of light relief, a little bit of fun, and just joking around. Funny apps on the phone. You know, where you change your voice, faces, all that kind of thing. It was just a little bit of laughter in those challenging times.’* participant 6, case two, nurse  **Organisational strategies**  *‘So then you come in to work and not being busy which didn't feel right, you felt almost a fraud coming to work and not having anything specifically to do with patients but sort of stepped up on staff wellbeing really so we developed what we call a wobble room and I put loads of resources in there for staff to do with, you know, breathing techniques and mindfulness.’* p*articipant 3 case four, Allied health care professional*  *‘I’ve spoken to him (chaplain), and he is really busy with staff members coming and talking to him about stuff. And I think at this kind of…at a time like this, spirituality is really important to a lot of people, and they will want to speak to someone about those kind of things, and so our chaplain’s finding that he’s giving himself…he’s offering a lending ear to some of these staff as well. So (inaudible 00:21:54) and listening to them has been really important, and we’ve kind of put that quite high on our priority list and we do have things in place.’* participant 2, case two, clinical manager  **Community strategies**  *‘We were very lucky in the community. We did a lot of appeals and a lot of schools and communities were donating PPE and we were just prioritising when to wear it. Obviously we had the masks which we were wearing all the time and our own visors. As the staff you look after the patient. But we had schools that were producing visors and people that were donating which was really, really helpful at the start because we just couldn’t order PPE at all. And there was always that sorry that we run out. But we didn’t luckily.’* participant 2, case five, nurse |
| **Moral comfort** |  | *‘when you kind of doubt things sometimes, you think oh God I wish I could have done more – I always say this to staff, just have a little read of some of the cards we have been sent through, during this time.  About the difference that actually we have made. We have gone not above and beyond, because it makes it sounds like you are doing something way above what you should but I know that our staff on that ward have done 100% what they could have done for our patients and relatives, in a really, really awful, difficult time.  And, I don’t doubt that, but that doesn’t make it any easier when you are thinking gosh if only it could have been this way – do you get what I mean?  I am aware that we have done what we could have done’*  Participant 5, case 1 , nurse |
